# USE OF THE NUCLEAR IRREGULARITY INDEX IN THE PROGRESSION OF ORAL CAVITY CARCINOMA: A STUDY MODEL

**DOI:** 10.1101/2025.04.07.25325404

**Authors:** Manuel García Flórez, Tatiana Carla Tomiosso, Fernando Bolaños

**Author notes:** Mail: Manuel García Flórez, Cellular Biology Laboratory, Faculty of Health, Universidad Surcolombiana, Calle 9 No 14-03, CP 400001, Neiva – Huila.

## Abstract

Oral cavity cancer, encompassing lip, mouth, and tongue cancers, is of interest because it is the sixteenth most prevalent neoplasia globally, accounting for 389,846 new cases and approximately 188,000 deaths by 2022.

The specific objective of this study was to investigate morphological changes in the nuclei of oral squamous cell carcinoma (OSCC) using specific DNA staining techniques and an automated Irregularity Nuclear Index (NII) for classification.

Samples from 32 patients diagnosed with OSCC were analyzed, with a focus on distinguishing tumor differentiation stages through nuclear morphometric analysis.

A mixed-method approach was employed using fluorescence microscopy and FIJI software to quantify nuclear irregularities, which revealed significant insights into apoptosis, mitosis, interphase, and senescence. These findings aim to enhance diagnostic accuracy and provide new tools for the early detection and management of oral cavity cancer. This research highlights the potential for integrating digital methodologies in pathology to bridge gaps in our understanding of the relationship between morphological nuclear changes and molecular alterations in tumor progression. Keywords: Squamous Cell Carcinoma of Head and Neck, Image Processing, Computer-Assisted, Apoptosis

## INTRODUCTION

Oral cavity cancer, including lip, mouth, and tongue cancer, represents the sixteenth most common neoplasia worldwide. At this scale, it was responsible for 355,000 new cases, and about 177,000 deaths in 2018 (Miranda-Filho & Bray, 2020). The geographical areas with most incidence are in the South and Southeast of Asia (India, Pakistan and Taiwan), in East Europe (Hungary, Slovakia and Slovenia), and parts of Latin America and the Caribbean (Warnakulasuriya, 2009).

Oral cavity cancer is conventionally diagnosed and classified based on its histopathological characteristics: morphological alterations identified in samples indicate the status and progression of the tumor. However, changes in the nuclear structure and texture that may precede histopathological changes, may sometimes not be visible to pathologists (Carleton, Lee, Madabhushi, & Veltri, 2018),(Damodaran, Crestani, Jokhun, & Shivashankar, 2019),(Fischer, 2020) and, although such changes alone are not the causes of cancer, they show the transformation and progress of the cells towards oncogenic events (Madrazo, Conde, & Redondo-Muñoz, 2017).

To manage it successfully, an early detection and proper approach is required, although high reappearance and aggressivity of the tumor decrease the treatment effectiveness (Liao et al., 2012),(Noble et al., 2016).

Bhargava, Damodaran and Fillippi-Chiela studies show different techniques, methodologies, and development of software that quantify changes in the nuclei of tumor cells in an automated way, leading areas like oncology and pathology towards a digital transformation, which have provided the specialists with new tools for obtaining a more accurate diagnosis (Bhargava & Madabhushi, 2016),(Damodaran et al., 2019),(Filippi-Chiela et al., 2012). However, there are still gaps in techniques and topics to be developed such as establishing a more direct relationship between morphological changes observed in nuclei and changes at the molecular level.

This study identifies, with a specific DNA staining, changes in the nuclei sizes of oral cavity cancer tumoral cells and, through the *Nuclear Irregularity Index* complement, they are automatedly classified into different groups. Thanks to this technique, morphological characteristics of apoptosis, mitosis, interphase, senescence, among others are visualized, and an association is established with the differentiation stages of the tumor, aiming at providing new tools of diagnostic utility.

### Patients and Methods

Paraffin-embedded samples were taken from patients diagnosed with oral squamous cell carcinoma (OSCC), coming from the Department of Pathology of Hospital Universitario Hernando Moncaleano Perdomo, Neiva – Huila.

The clinical observation period was 36 months from 2014 until 2016. Samples were selected from 32 patients, and after pathology assessment, they were classified according to the tumor differentiation stage as: well differentiated, moderately differentiated, and poorly differentiated, highlighting the tumor infiltration and keratinization (Table 1).

**Table 1.** Summary of the main characteristics found in the one convenience selected sample of patients with oral cavity squamous cell carcinoma. Tumor differentiating degree, sample numbers and injury location are shown.

| Degree of differentiation of SCC*. | n | Anatomic Location |
| --- | --- | --- |
| Well differentiated | 16 | Tongue (12), Tonsil (1), Larynx (3) |
| Moderately differentiated | 13 | Tongue (7), Tonsil (1), Larynx (3), Lip (2) |
| Poorly differentiated | 3 | Tongue (1), Tonsil (1), Lip (1) |

SCC=squamous cell carcinoma

### Fluorescence microscopy

DAPI staining, (Abcam ab104139), specific for identifying DNA, was used to evidence nuclear morphology. For that, plates were deparaffinated and washed with two refrigerated xylol 1-min repetitions (Sigma, Aldrich), then three 95 % ethanol changes, and three changes of TBS buffered saline solution were made.

DAPI was then applied (1 µg/mL) for 5 min, followed by two TBS changes, later covered with microscope slide, and taken to a fluorescence lamp-equipped microscope. Images were captured with AxioCam ICm1 (Carl Zeiss, Göttingen, Germany) coupled to the Axio imager Z2 Microscope with focus on motorized z, and obtained with the objective EC Plan-Neofluar 40x/0.75 M27 (Carl Zeiss, Göttingen, Germany). The Set 49 set of filters was used, which detects 358 nm excitation, and emission at 463 nm.

Images were captured with 300 dpi resolution and in TIFF format with the ZEN Digital Imaging® (Carl Zeiss, Göttingen, Germany).

### Nuclear Morphometric analysis

Analysis of nuclear morphology was made in FIJI program (Schindelin et al., 2012), freely available on https://imagej.net/software/fiji/downloads, in which the complement *Nuclear irregularity Index (****NII***) was installed, developed by Filippi-Chiela (Filippi-Chiela et al., 2012). FIJI allows obtaining morphometric information of nuclei by quantifying the nuclear area and four irregularity parameters to generate the **NII (Figure 1)**, from which data on the morphological characteristics of nuclei and the biological significance of the different cellular populations are obtained (Table 2).

**Table 2.** Morphological characteristics and biological meaning of the different cell populations of the morphometric nuclear analysis shown in Figure 4. Taken and adapted from Filippi-Chiela et al., (Filippi-Chiela et al., 2012).

| Symbol | Name | Location in graph | Morphological appearance of the nucleus | Cellular significance |
| --- | --- | --- | --- | --- |
| N | Normal | Nucleus within or close to the ellipse defined by a population of normal/regular nuclei | Normal size and shape | Interphase without alterations that affect the nuclear morphology |
| I | Irregular | Nucleus with area similar to nucleus N, but with high NII | Normal size and high irregularity | Catastrophic mitosis or other event that affects nuclear morphology |
| SR | Small regular | Nucleus with area smaller than the normal ellipse, with low NII | Condensed and regular | Apoptosis in initial or intermediate stages |
| S | Small | Nucleus with area smaller than the normal ellipse, with intermediate NII | Small, but not spheric | Mitosis |
| SI | Small and irregular | Nucleus with area smaller than the normal ellipse, with high NII | Condensed, small and irregular | Mitosis with damages or nuclear fragments |
| LR | Big regular | Nucleus with area greater than the normal ellipse, with NII similar to N | Big and regular | Senescence |
| LI | Big and irregular | Nucleus with area higher to the normal ellipse, with high NII | Big with damage or multinucleated nucleus | Catastrophic mitosis or another event that affects the nucleus |

**Figure 1.**
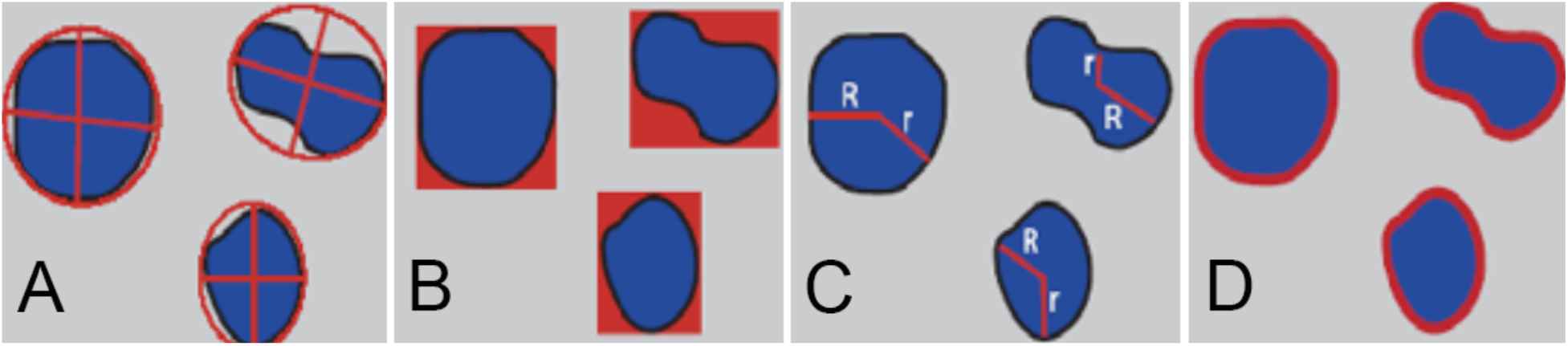
Illustration of four parameters that represent the observed variability in nuclear morphology and are employed to calculate an index referred to as the Nuclear Irregularity Index (NII). **Fig 1A**, Aspect (**Asp**) represents the ratio between the major axis and the minor axis of the ellipse that is equivalent to the object. It is noteworthy that the aspect is consistently greater than 1; **Fig 1B**, Area Box (**Arbx**) signifies the ratio between the area of each object and the area of its hypothetical bounding box, which is determined by the division of the area of the object by the area of the box; **Fig 1C**, Radius ratio (**Rr**) illustrates the ratio between the maximum radius and the minimum radius of each object; **Fig 1D**, presents the Roundness (**Rou**) of each object, which is ascertained by the following formula: (per2 1/(4* pi*area). It is crucial to note that circular objects will possess a roundness value of 1, whereas other shapes will have a roundness value greater than 1. Therefore, the NII is given by: NII= Asp-Arbx+Rr+Rou Taken and adapted from Filippi-Chiela et al., (Filippi-Chiela et al., 2012).

### Sample selection and statistical analysis

At least three different fields were randomly selected in the different groups of tumoral samples, counting between 300 and 500 nuclei approximately by experimental group.

Results are shown as percentages of the different classified groups. Likewise, the average and standard deviation for each of them are presented (Table 3).

**Table 3.** Nuclear characteristics grouped in the different stages of tumoral progression; well differentiated, moderately differentiated, and poorly differentiated. The percentage of normal nuclei (N), small nuclei (S), small regular (SR), small irregular (SI), large regular (LR), large irregular (LI), and irregular nuclei (I) is shown.

| Squamous Cell Carcinoma | N | S | SR | SI | LR | LI | I | Total number of nuclei |
| --- | --- | --- | --- | --- | --- | --- | --- | --- |
| Well differentiated | 63% (272) | 2% (8) | 12% (52) | 1% (4) | 10 (43) | 2% (10) | 10% (43) | 431 |
| Well differentiated, ulcerated and infiltrating | 56% (266) | 5% (22) | 20% (96) | 1% (6) | 1% (5) | 1% (7) | 19% (88) | 473 |
| Well differentiated, keratinizing and infiltrating | 13.7% (67) | 13.9% (68) | 65.5% (321) | 4.3% (21) | 1% (5) | - | 1.6% (8) | 490 |
| Well differentiated, keratinizing, and infiltrating of big cells | 17.7% (55) | 15.8% (49) | 53.1% (165) | 3.5% (11) | 1.0% (3) | - | 9% (28) | 311 |
| Well differentiated, keratinizing, and infiltrating of endophytic growth, ulcerated | 38.6% (117) | 5.3% (16) | 47.2% (143) | 0.3% (1) | 2.6% (8) | - | 5.9% (18) | 303 |
| Moderately differentiated | 11.2% (40) | 10.4% (37) | 71.9% (256) | 2% (7) | - | - | 4.5% (16) | 356 |
| Moderately differentiated, ulcerated, and infiltrated | 29.7% (148) | 9.8% (49) | 48.8% (243) | 1.8% (9) | 1.2% (6) | 2.2% (11) | 6.4% (32) | 498 |
| Poorly differentiated, infiltrating | 9.9% (38) | 18.8% (72) | 58.5% (224) | 7.3% (28) | - | - | 5.5% (21) | 383 |
| Average | 125.4 | 40.1 | 187.5 | 10.9 | 8.8 | 3.5 | 31.8 |  |
| DS | 96.5 | 23.6 | 89.6 | 9.1 | 14.1 | 5.0 | 25.1 |  |
| n | 1003 | 321 | 1500 | 87 | 70 | 28 | 254 |  |

## Results

The histopathologic analysis allowed classifying the samples in the different stages of tumoral progression from well differentiated, through moderately differentiated until poorly differentiated.

The microscopic characteristics of the well differentiated squamous cell carcinoma reveal irregular cells and some accumulations of keratin (keratin beads) (Fig. 2B), in the moderately differentiated squamous cell carcinoma the loss of cellular cohesion stands out (Fig. 2C), and in the poorly differentiated squamous cell carcinoma the cellular epithelial pattern is lost and various abnormalities such as hyperchromasia and pleomorphism appear (Fig. 2D).

**Figure 2.**
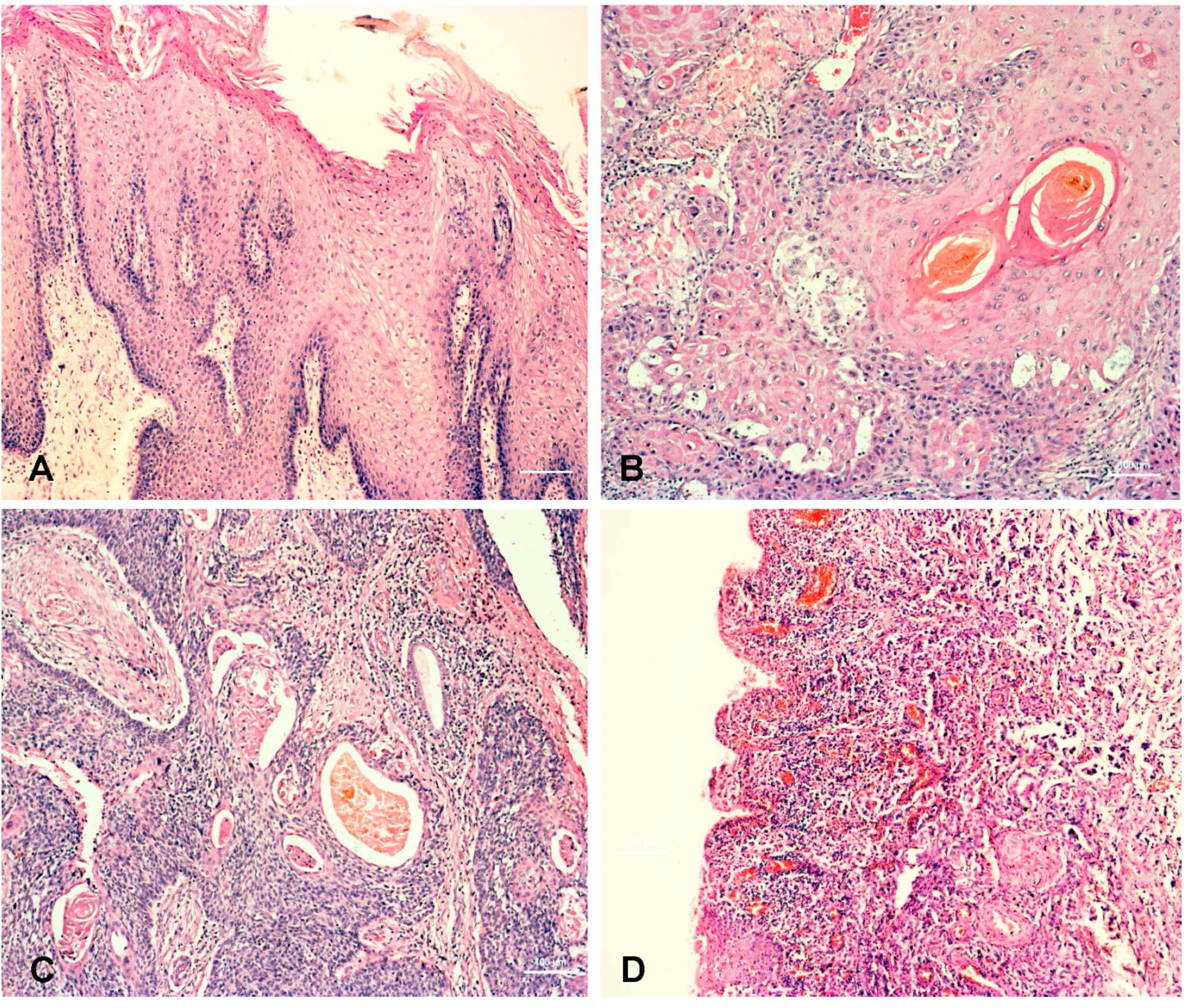
Tissue samples of the experimental groups used in the staining study with Hematoxylin and Eosin in low magnification (10X). 1A. Section of the tongue normal tissue, evidencing the normal stratified epithelium. Nuclei appear with regular shape and uniformly distributed. 1B. Well differentiated squamous cell carcinoma with keratin injury (keratin bead). Loss of stratification and irregularity in the nuclei distribution in tissue are observed. 1C. Moderately differentiated squamous cell carcinoma. Inflammatory cells infiltration and tissue irregularity are observed. 1D. Poorly differentiated squamous cell carcinoma. Total loss of the tissue organization is evident. Cell dedifferentiation showed an increase in the number of nuclei, accompanying their area reduction. Bar 100 microns

Coloration with DAPI allowed identifying the nuclei in the different treatments. In the samples used as control, regular and uniform nuclei were identified. Several characteristics of the nuclei were detected and organized according to their regularity and size (Fig.3). Based on these data, the nuclei of the other experimental groups were compared generating values, a distribution graph, and a table with the results (Table 3). The control group shows nuclei with a regularity index (**NII**) approximated to 1 and with an approximate average area of 1500 pixels^2^ (Figure 4, ellipse located in the lower left quadrant). The nuclei from the other experimental groups are observed as small rhombuses and squares located in the graph. It is remarkable that a large part of these nuclei, although keep a regularity index close to 1, show a reduction in their area. Others, however, although in smaller quantities, show higher areas and a regularity index greater than 1.

**Figure 3.**
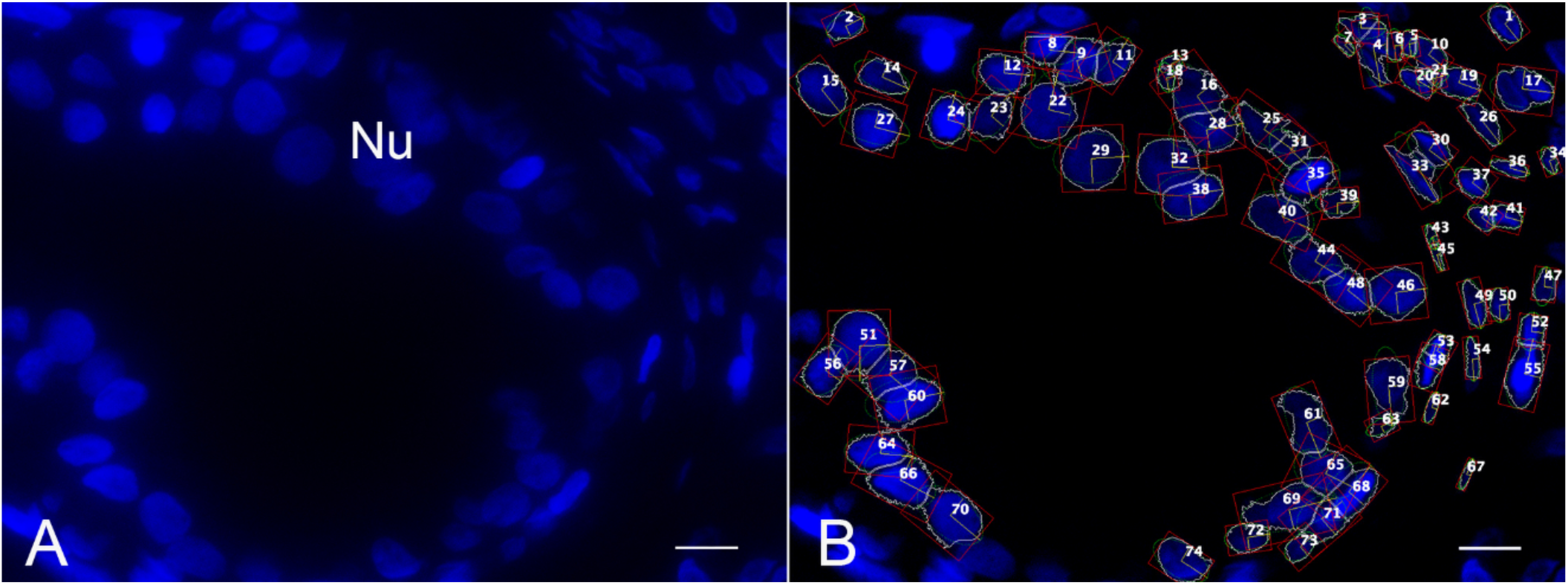
Process of visualization, selection, and quantification of the characteristics of the cellular nuclei in the different experimental groups. 2A. Image capture of representative areas in the different treatments in slides previously treated with DAPI to evidence the nuclear DNA. Selection and standardization of nuclei to prevent their over registration and under registration. 2B. Performing of the Nuclear Irregularity Index (NII) complement of FIJI software that yields nuclear morphometric information. Bar 10 microns.

**Figure 4.**
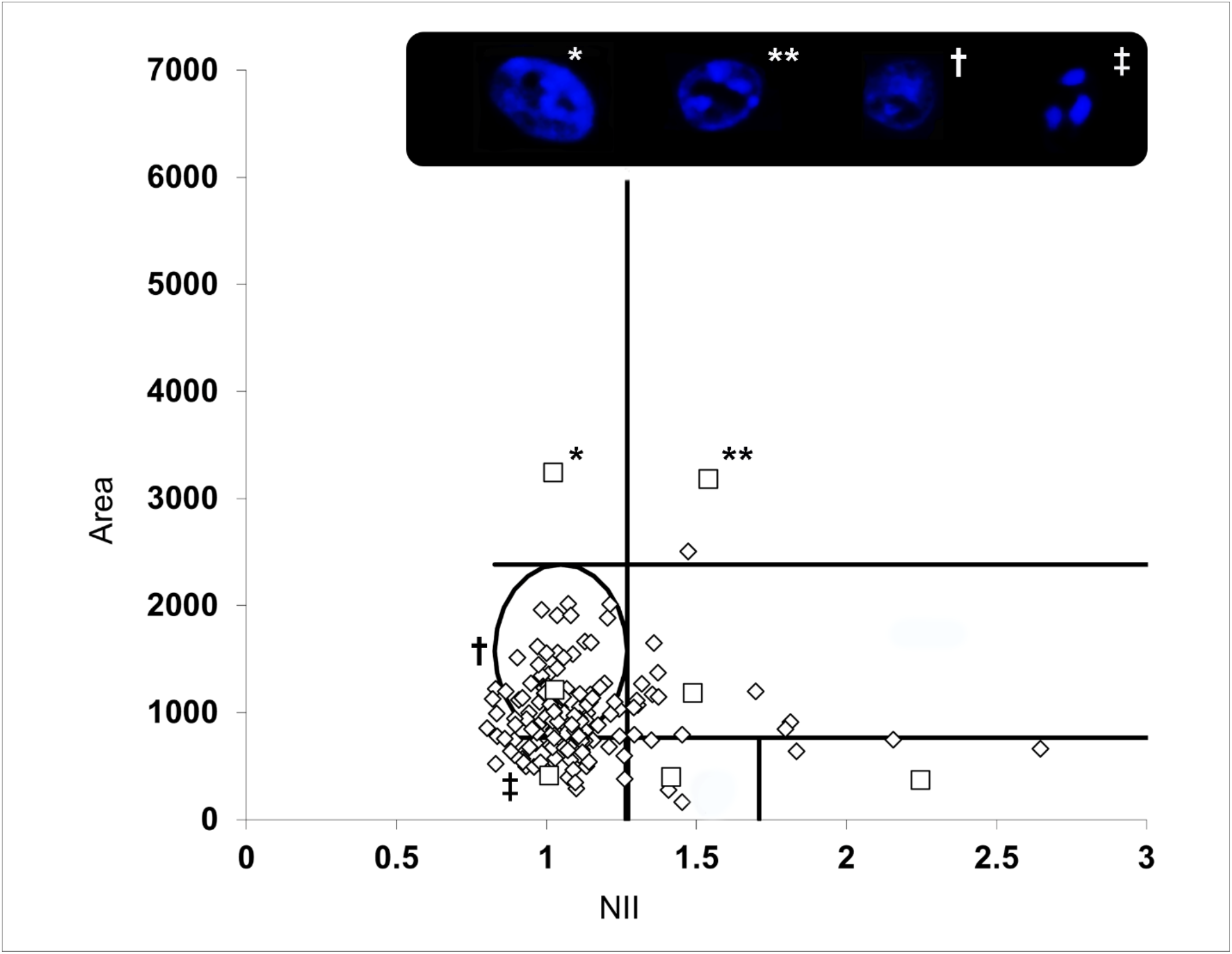
Nuclei distribution graph of area vs. Nuclear Irregularity Index (NII). Normal nuclei are shown, used to establish the reference population, represented by those within the ellipse. Those out, represent the different nuclei populations. Photos and symbols show nuclei examples and their location in the graph. **\***: Big regular; **\*\***: Big and irregular; **†**: Normal; **‡**: Small and irregular.

When taking those data and grouping them according to the histopathological classification of well, moderately, and poorly differentiated, it was found that cells from well differentiated squamous cell carcinoma have mainly nuclei in interphase without alterations that affect nuclear morphology, followed by nuclei in initial or intermediate stages of apoptosis and nuclei in senescence stage. Instead, when the tumor is keratinized and infiltrating, nuclei in initial or intermediate apoptosis increase, and reach over half of the nuclear population.

It is also noticeable that in moderately differentiated cancer nuclei in interphase without alterations fall dramatically to 10 % or 30 % of the nuclear population. The rest of the nuclei percentage are not homogeneous and are distributed in the different groups, but with a higher proportion of apoptotic nuclei. Poorly differentiated cancer has approximately 10 % of nuclei without morphological alterations or in interphase, senescent and in catastrophic mitosis nuclei are scarce or little detectable, but by adding the group of small nuclei, a 90 % of the population is obtained: of these, in descending order, nuclei in apoptosis, nuclei in mitosis, and nuclei in mitosis with damages of nuclear fragments were found (Table 3).

## Discussion

Although there are different high-precision genomic tests that recognize molecular changes occurring during cancer progression, they sometimes do not allow detecting the heterogeneity of the disease because generally a small sample of a tumoral area is analyzed. A solution could be to make an image analysis supported by specific software, to quantify a considerable number of images covering tumoral and benign areas to identify variations in the nuclear morphology of cells (Carleton et al., 2018).

Similar image analysis techniques have been used to examine the nucleus structure in cases of localized ductal breast cancer (Tan, Goh, Chiang, & Bay, 2001),(Irshad, Veillard, Roux, & Racoceanu, 2014),(Hayward et al., 2020) and oral squamous cell cancer (Padovani et al., 2007), (Kumar, Chatterjee, Purkait, & Samaddar, 2017), (Natarajan, Mahajan, Boaz, & George, 2009).

However, all these reports show the implementation of this technique using general dyes such as hematoxylin and eosin (H&E) staining. Although these dyes offer a comprehensive overview and serve as standard stains in the field of pathology, they do not provide the precision required to deeply analyze nuclear morphology. Our study, on the other hand, offers an advantage by utilizing 4’,6-Diamidino-2-phenylindole (DAPI) staining, which exhibits greater specificity through selective binding to the minor groove of DNA. Furthermore, DAPI staining possesses lower mutagenic properties and does not present the carcinogenic and/or the toxicity potential of other fluorescent dyes like propidium iodide or Hoechst 33342 (Atale, Gupta, Yadav, & Rani, 2014).

The found results match previous reports, where the decrease of the nuclear area is related to the degree of differentiation of the tumor. Initially, the cell and nuclear volume increase, but as the progression of dysplastic stages occurs towards carcinomas, a reduction of the cellular radius occurs (Vedam, Boaz, & Natarajan, 2014).

On the other hand, it has been established that alterations of the balance between cellular proliferation and death appear in cancer, where the rate of proliferating mutant cells surpass the number of lost cells. In normal tissues, apoptosis is a control mechanism of the cellular population expansion, keeps equilibrium and eliminates cells affected by potential damages in their DNA.

However, in poor prognosis cancer, besides high proliferation rates, it is also possible to find high indexes of apoptosis (Morana, Wood, & Gregory, 2022), which has been verified in other types of neoplasia such as bladder cancer (Jalalinadoushan, Peivareh, & Azizzadeh Delshad, 2004), breast cancer (Villar et al., 2001), colorectal cancer (Alcaide et al., 2013), gastric cancer (Koshida, Saegusa, & Okayasu, 1997), glioblastoma (Sarkar et al., 2005), and squamous cell carcinoma of the tongue (Naresh, Lakshminarayanan, Pai, & Borges, 2001), among others.

Morana et al., have assumed that programmed cellular death can produce pro-oncogenic signals, where a cell population can lead to the alteration of the proliferation/death equilibrium, one of the key factors of carcinogenesis (Morana et al., 2022).

Different ways of cellular signaling can be activated, and a particularly interesting way is that of Tumor-associated macrophages (TAMs), cells that, together with others, take charge of recycling the apoptotic cellular remainder and its accumulation in tissue. TAMs would produce pro-oncogenic signals that, in some types of cancer, would induce the tumoral growth and the angiogenesis process (Mantovani, Sozzani, Locati, Allavena, & Sica, 2002),(Sica et al., 2008).

## Conclusions

Despite the existence of different specific molecular tests to detect different cellular processes such as apoptosis, the heterogeneity of cancer brings with it a challenge that always requires new instruments. Specific software-aided analyses are a tool that is becoming more and more relevant every day in different analyses of tissues or structures of biological interest. Their use in pathology could be oriented as a tool for a quick analysis of large quantity of images to obtain qualitative results that allow orienting diagnoses or elucidate signaling ways of different pathological processes. For instance, the most relevant finding of our study was the high rates of cell death in poorly differentiated cancer samples (approximately 60%). This finding was not expected, but this feature has been previously reported in other types of tumors and has been linked to tumor aggressiveness (Morana et al., 2022).

## Data Availability

All data produced in the present study are available upon reasonable request to the authors

## Acknowledgments

The authors thank the patients and the Pathology Service of Hospital Universitario Hernando Moncaleano Perdomo for the loan of samples that allowed making this research.

## Financial Support

The experiments conducted had the support of the vice rectory of research and social projection of Universidad Surcolombiana.

## Conflict of Interest

The authors declare that there is no conflict of interest about the making of this project.

## Authors contributions

FB: Carried out the tumors classification and histopathological analysis and contributed to the final manuscript.

TCT: Review and write of the final manuscript.

MGF: Carried out the experimental design, identification, and classification of the nuclei by fluorescence, image capture and analysis, composition and editing of the final manuscript.

## Notes

### Competing Interest Statement

The authors have declared no competing interest.

### Funding Statement

This study was funded by Vicerrectora de Investigaciones Universidad Surcolombiana

### Author Declarations

Ethics committee/IRB of Hospital Universitario Hernando Moncaleno Perdomo gave ethical approval for this work. Code number 003-003

